# DNA Methylation Differences Stratified by Normalized Fetal/Placental Weight Ratios Suggest Neurodevelopmental Deficits in Neonates with Congenital Heart Disease

**DOI:** 10.1101/2025.01.08.25320236

**Authors:** Marin Jacobwitz, Michael Xie, Jamie Catalano, Ingo Helbig, J. William Gaynor, Nancy Burnham, Rebecca L. Linn, Juliana Gebb, Mark W. Russell, Barbara H. Chaiyachati, Ana G. Cristancho

## Abstract

**Background:** We lack early biomarkers for predicting neurodevelopment (ND) outcomes in children with congenital heart disease (CHD). Placentas of fetuses with CHD have abnormalities, including unbalanced fetal/placental weight ratios (F/P). Although DNA methylation profiles have revealed insights into the maternal-fetal environment (MFE), it is unknown if DNA methylation correlates to normalized F/P weight ratio groups and how these differences relate to ND outcomes.

**Methods:** We prospectively recruited a cohort of pregnant women carrying a fetus with CHD. A subset of the cohort had DNA methylation performed on either umbilical cord blood or postnatal blood (45 full-term neonates). We calculated normalized F/P weight ratios, focusing on three normalized F/P ratio groups for analysis. We calculated differential methylation signals in eight ND disabilities-associated gene sets. Normalized F/P ratios were compared to 18-month Bayley Scales of Infant Development-III scores (BSID-III).

**Results:** Unbiased gene ontology enrichment analysis of differentially methylated regions revealed enrichment for brain development-related pathways. Although there were no significant differences between normalized F/P weight ratio groups and BSID-III, disease-associated gene set pathway analysis revealed significant methylation differences between the most severely unbalanced F/P weight ratio and normal F/P weight ratio groups.

**Conclusion:** Gene ontology enrichment analysis of differential methylation regions revealed significant differences between normalized F/P weight ratio groups in neurogenesis genes. Furthermore, our data identified methylation differences between unbalanced and balanced normalized F/P weight ratio groups in gene pathways associated with ND dysfunction common in the aging CHD population suggesting converging pathways for ND disorders that should be investigated further.

## Introduction

Congenital heart disease (CHD) is the most common birth defect, impacting approximately nine per 1,000 live births, with three per 1,000 having critical CHD requiring catheter-based intervention or surgery in the neonatal period to ensure survival.^1,2^ With improvements in surgical and medical outcomes, children with CHD are now surviving well into adulthood, which has allowed for longitudinal neurodevelopmental surveillance.^3,4^ It has become apparent that many survivors of CHD have an array of neurodevelopmental disabilities impacting their quality of life and adult independence, including autism, visuospatial deficits, attention deficit-hyperactivity disorder (ADHD), and epilepsy.^3–8^ Genetic predisposition may account for some, but not all, of the adverse neurodevelopmental outcomes, as only 25-40% of children with CHD have extracardiac anomalies or an identified genetic syndrome,^9^ and the currently known postnatal, medical, and surgical risk factors only account for approximately 30% of the variation in these long-term outcomes.^10,11^ Although earlier theories focused on neonatal surgery as the primary culprit contributing to adverse neurodevelopmental outcomes, recent research has identified that an adverse in-utero environment, including abnormal placentology, may be the foundation of brain vulnerability with additive insults postnatally.^12–15^

The placenta is a highly vascular transient organ that acts as an interface for oxygen and nutrient delivery from the mother to the fetus.^16,17^ The placenta has been increasingly implicated in adverse neurodevelopmental outcomes in CHD, with many studies identifying high rates of vascular and structural abnormalities in postnatal placental pathology in CHD pregnancies.^16,18–20^ The severity of placental abnormalities in postnatal pathology has been correlated with smaller neonatal brain volumes, echoing the role of the placenta in fetal brain growth.^18^ Neonates with CHD and superimposed placental dysfunction from gestational hypertension, preeclampsia, growth restriction, and pre-term birth demonstrated increased length of hospital stay and higher mortality than neonates with CHD and healthy placentas.^21^ One theory is that the placenta in CHD has a structurally abnormal phenotype, leading to the inadequate oxygenation of fetal blood, with subsequent lower oxygen saturation of blood from the placenta.^22^ In the already vulnerable fetus with CHD, this theory results in lower cerebral oxygen delivery, poor brain development, and ultimately impaired neurodevelopment.^22^ With ongoing evidence that many survivors of CHD have neurodevelopmental disabilities regardless of cardiac lesion type, investigations into the influence of the maternal-fetal environment (MFE) and placenta on longitudinal neurodevelopmental outcomes are increasingly being investigated.^23,24^

Categorization of fetal to placental ratio (hereafter F/P) into either “balanced,” which is normal growth of the fetus and the placenta relative to one another, or “unbalanced,” describing discrepant growth of the fetus and placenta, can be used in lieu of merely evaluating the growth of the fetus or the placenta as separate entities. For example, using a 9-block categorization based on placental weight and F/P weight ratio z-scores to create normalized F/P weight ratios, previous research has identified that there was an increased risk of perinatal death with unbalanced F/P growth compared to balanced F/P growth.^25^ Furthermore, using placental weight and F/P weight ratio z-scores to classify normalized F/P weight ratios into clinically relevant categories allows for a better understanding of how the fetus and the placenta are growing relative to one another and, therefore, the health of the pregnancy.^25^

To specifically understand the importance of normalized F/P weight ratios, here we focus on the relationship between normalized F/P weight ratios and the fetal and neonatal DNA methylation profile. The health of the placenta likely impacts the epigenetic landscape of the fetus and newborn infant.^26,27^ Epigenetic regulation of gene expression is a well-regulated process necessary for normal development. Dysregulation of the epigenome can result in pathology, including neurodevelopmental disorders.^28–30^ Specifically, DNA methylation has been identified as an important part of epigenetic regulation in the placenta, with dynamic changes throughout gestation that may influence fetal and placental growth and function.^27,29–32^ The objective of this paper was to identify if there are differences in DNA methylation of fetal or infant blood between balanced and unbalanced normalized F/P weight ratio groups in a cohort of neonates with CHD. Gene pathways associated with common neurodevelopmental dysfunction identified in survivors of CHD were specifically examined to determine if there may be a clinical relevance to these normalized F/P weight ratios and relevant outcomes.

## Methods

### Subject Cohort

The cohort was prospectively recruited for a study focusing on environmental exposure influence on neurodevelopmental outcomes in neonates with CHD and has been previously published (hereafter referred to as “parent study”).^11^ Written informed consent was obtained from all pregnant patients, and the study was approved by the Institutional Review Board at The Children’s Hospital of Philadelphia. As part of the parent study, umbilical cord blood was collected from the neonates at the time of birth. If umbilical cord blood was unable to be collected, a postnatal blood sample was obtained. A subset of the cohort had DNA methylation profiles performed using Infinium MethylationEPIC BeadChip V1 (850K) microarray^33^ in the Center for Applied Genomics at The Children’s Hospital of Philadelphia following the standard protocol from Illumina. Neurodevelopmental testing was completed at 18 months of age using The Bayley Scales of Infant and Toddler Development-III (BSID-III), which provide composite scores for motor, language, and cognitive skills, with higher scores indicating better skills.^34^ For statistical analysis purposes, the neurodevelopmental scores were categorized by sex assigned at birth. Data for this study was accessed on April 4, 2023, for research purposes. For this study, the authors did not have access to information that could identify individual participants during or after data collection.

### Inclusion/Exclusion Criteria

Inclusion criteria for the parent study were infants with CHD and expected cardiopulmonary bypass surgery at age 44 weeks post-conception or younger.^11^ Subjects with an identified genetic syndrome, major extracardiac anomaly, or language other than English spoken in the home were excluded.^11^ A total of 988 pregnant women were screened for eligibility in the parent study, with 848 subjects excluded for not meeting parent study inclusion criteria (700), declined to participate in the parent study (130), and other reasons (18) in the parent study.^11^ 140 pregnant women were enrolled in the parent study, with 110 infants (79%) returning at 18 months for neurodevelopmental (ND) follow-up.

Additional inclusion and exclusion criteria were applied for this study based on principal component and methylation age analyses. Reported race and ethnicity have been shown to be associated with differential DNA methylation patterns, including differences in methylation in cord blood,^35^ consistent with our principal component analysis (Supplemental Figure 1). We therefore excluded subjects that did not identify as non-Hispanic white (N = 31) as we were underpowered for downstream analysis. Of the remaining 79 subjects, 12 subjects did not have DNA methylation profiles and were therefore excluded. Given the known impact of maternal co-morbidities^36,37^ and cigarette smoking^38^ on DNA methylation, all subjects with an impaired maternal-fetal environment (MFE), defined as pre-existing maternal comorbidities, gestational comorbidities, and cigarette use, were excluded (N = 15). All subjects with samples collected >30 days postnatally were excluded (N = 1). Of the remaining 51 subjects, 2 subjects were not within the three F/P weight ratio groups of interest, and 4 subjects failed quality control. Our final cohort consisted of 45 subjects (Figure 1). A comparison of umbilical cord blood and postnatal neonatal blood DNA samples of the 45 subjects revealed no significant differences in cell type composition (Supplemental Figure 2); therefore, we included subjects that had either umbilical cord blood or postnatal DNA samples. Samples with a mismatch between sex determined by their methylation array data and self-reported sex were also excluded.^33–36^

**Figure 1.**
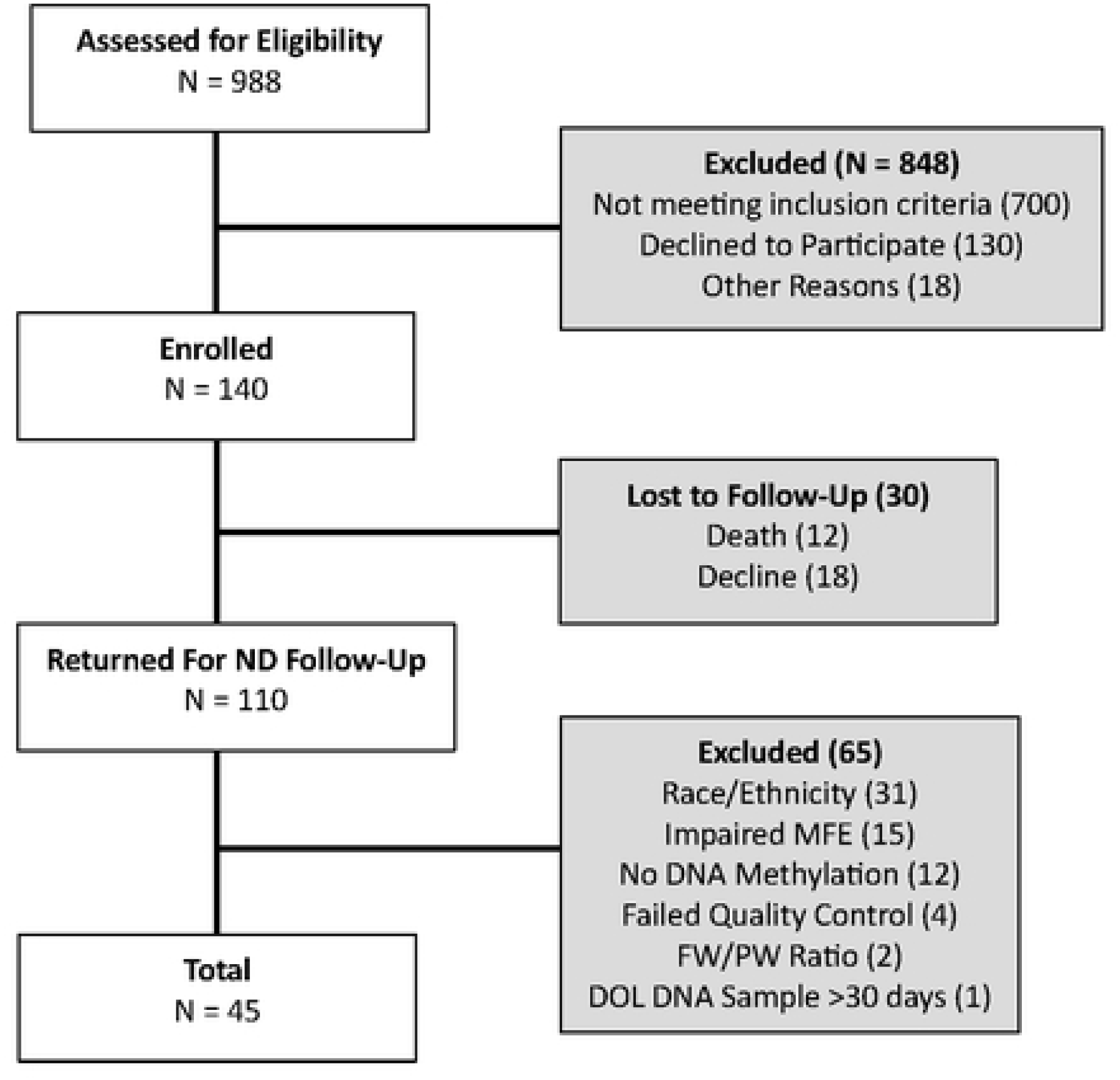
Cohort flow diagram

### Fetal/Placental Weight Ratio Categorization

The normalized fetal/placental (F/P) weight ratios were calculated for all subjects using the method previously reported by Matsuda et al. (2018).^25^ Matsuda et al. (2018) utilized placental weight and F/P weight ratio z-scores to establish a 9-block categorization of normalized F/P ratios to better understand F/P growth and perinatal outcomes.^25^ Subjects with normalized F/P weight ratios that were one of the following three categories were of interest and therefore included in this study: inappropriately light placenta with a relatively heavy infant (Group A), light placenta with relatively balanced growth of infant (Group D), and balanced growth of both placenta and infant (Group E).^25^ All other F/P categories were excluded due to sample size (Group F, N = 1; Group B, N = 1).

### Infinium MethylationEpic BeadChip Data Processing and Quality Control

Minfi R package (Version 3.18, ref [14], R version 4.3) was the primary data analysis pipeline. Raw idat files were imported and digitalized using “read.metharray.exp” function with all default parameters. We removed any poor-quality probes (detection p-value ≥ 0.01) and any sample with poor-quality probe rate (more than 5% of total probes). We removed cross-reactive probes.^39^ Sex chromosomes were removed from the analysis to reduce sex-related variation given the small sample size. Quantile normalization method (“preprocessQuantile” function) was then applied to minimize unwanted variation between samples and reduce potential batch effects. The beta value was calculated using “getBeta” function and then logit transformed to M value using the “getM” function. M values for 683083 CpG probes were retained for follow-up statistical analysis.

### DNA Methylation Differential Analysis

After initial quality control and export of M values after normalization, the association between the three normalized F/P weight ratio categories, as described above, and probe methylation were evaluated. Linear models were fit for each probe (“lmFit” function in limma, version 3.54) with “sex” (“M” or “F”) and biospecimen source (“umbilical cord blood” or “whole blood DNA”) as covariates. All test statistics were modified by empirical Bayes procedure to protect against test statistic inflation.^40^ Each contrast was studied and reported separately.

### Gene Set Enrichment Analysis

Based on results from differential analysis, a list of methylation sites with p-value < 0.05 was generated. The functional enrichment characteristics among those sites were evaluated by Gene Ontology annotation within Illumina Epic Array (“gometh” function in missMethyl, version 1.24). Additionally, eight gene sets were targeted based on commonly identified neurodevelopmental disabilities previously reported in the CHD population:^3,5,6,8,41–43^ “Alzheimer’s,”^44^ “autism,”^45^ "epilepsy,”^46^ “Attention Deficit Hyperactive Disorder (ADHD),”^47 40^"executive dysfunction,"^47^ "social anxiety,”^47^ "visuospatial dysfunction,”^47^ and “positive regulation of angiogenesis”^48,49^ gene sets. We utilized “methylgometh” function (“methylGSA” package, version 1.8) to measure gene set enrichment among our methylation sites. “methylgometh” function adjusts the number of CpGs for each gene by weighted resampling and Wallenius non-central hypergeometric approximation. We report p-value and Benjamini-Hochberg false discovery rate (FDR) adjusted p-values for each targeted gene set we curated for this study.

### Placental Groups and BSID-III

The cohort was first stratified by sex, as neurodevelopmental outcomes demonstrate sex differences.^50^ We then examined the differences between the normalized F/P weight ratio groups and the BSID-III for language, motor, and cognition. A one-way ANOVA test (aov function, R stats version 3.6.2) was employed to test the significance of the association. Each comparison was tested and reported separately.

## Results

Demographic and clinical characteristics of the cohort are in Table 1. Forty-nine percent (22/45) of the cohort was male (Group A 4/11, 36%; Group D 5/11, 45%; Group E 13/23, 57%). The most common cardiac lesion was transposition of the great arteries (TGA) in 38% (17/45) of subjects, followed by 33% (15/45) with hypoplastic left heart syndrome (HLHS). The average gestational age (GA) at birth was 39.23 weeks (SD 0.75 weeks). Forty percent (18/45) of the cohort had methylation profiles on umbilical cord blood sample; the remaining 60% had methylation microarrays performed on postnatal peripheral blood sample. Average day of life collection of blood samples was 5.7 days (SD 5.3 days; Table 1).

**Table 1.** Demographics.

| Variable<br>Mean ± SD | Cohort<br>N = 45 (%) | Group A<br>N = 11 (%) | Group D<br>N = 11 (%) | Group E<br>N = 23 (%) | p value |
| --- | --- | --- | --- | --- | --- |
| Sex - male | 22/45 (49) | 4/11 (36) | 5/11 (45) | 13/23 (57) | 0.5276* |
| Gestational Age at Birth | 39.23 ± 0.75 | 39.52 ± 0.50 | 39.17 ± 0.77 | 39.12 ± 0.82 | 0.148** |
| Cardiac Lesion Type |  |  |  |  | 0.97* |
| HLHS | 15/45 (33) | 4/11 (36) | 3/11 (27) | 8/23 (35) |  |
| TGA | 17/45 (38) | 4/11 (36) | 5/11 (45) | 8/23 (35) |  |
| Other | 12/45 (27) | 3/11 (27) | 3/11 (27) | 6/23 (26) |  |
| Sample Type |  |  |  |  | 0.7658* |
| Cord Blood | 18/45 (40) | 5/11 (45) | 5/11 (45) | 8/23 (35) |  |
| Postnatal DNA | 27/45 (60) | 6/11 (55) | 6/11 (55) | 15/23 (65) |  |
| DOL DNA Sample | 5.7 ± 5.3 | 3.18 ± 4.81 | 2.36 ± 2.54 | 4.04 ± 5.93 | 0.688** |
| *Pearson's chi-squared test<br>**one-way ANOVA test |  |  |  |  |  |

### Placental Groups and Gene Ontology Enrichment Analysis

An unbiased gene ontology enrichment analysis of differential methylation regions revealed enrichment for pathways related to brain development and neuronal function. Normalized F/P weight ratio Group A (inappropriately light placenta with relatively heavy fetus) and normalized weight ratio Group E (normal) were enriched for differential methylation signals in the following pathways: *nervous system development* (GO:0007399), *neuron differentiation* (GO: 0030182), *generation of neurons* (GO:0048699), *neuron development* (GO:0048666), *synapse* (GO:0007399), and *neuron projection* (GO:0043005; Figure 2); normalized F/P weight ratio Group D (light placenta with relatively balanced growth of fetus) and normalized F/P weight ratio Group E (normal) were enriched for differential methylation signals in the following pathways: *neurogenesis* (GO:0022008), *regulation of neuron projection development* (GO:0010975), *neuron projection development* (GO:0031175), *neuron development* (GO:0048666), and *generation of neurons* (GO:0048699; Figure 2); normalized F/P weight ratio Group A (inappropriately light placenta with relatively heavy fetus) and normalized ratio Group D (light placenta with relatively balanced growth of fetus) were enriched for differential methylation signals in the *nervous system development* (GO:0007399) and *neurogenesis* (GO:0022008; Figure 2) pathways.

**Figure 2.**
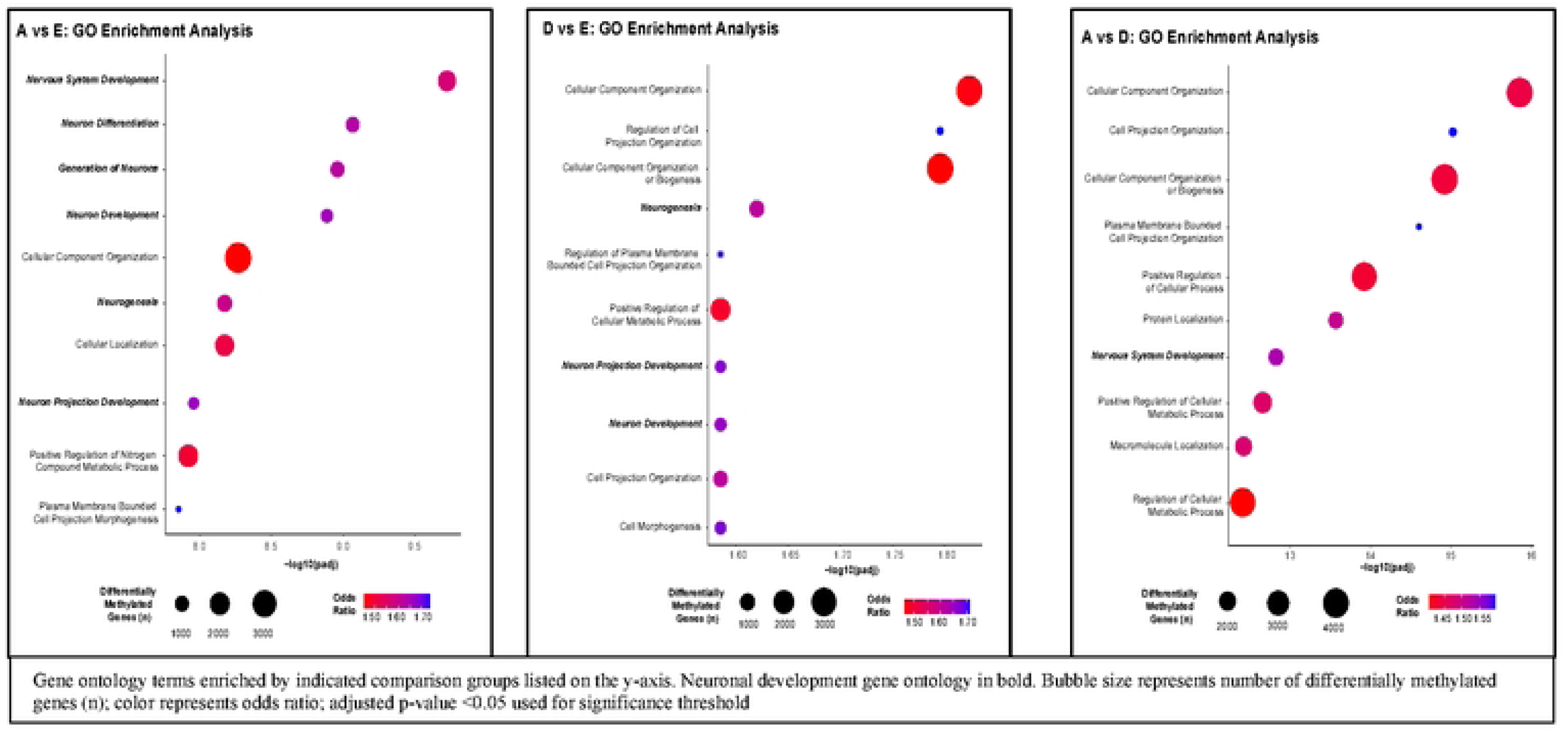
Placental Groups and Gene Ontology Enrichment Analysis, Gene ontology terms enriched by indicated comparison groups listed on the y-axis. Neuronal development gene ontology in bold. Bubble size represents the number of differentially methylated genes (n); color represents odds ratio. We report p-value and Benjamini-Hochberg false discovery rate (FDR) adjusted p-values for each targeted gene set.

### Placental Groups and ND-associated Gene Pathways

When comparing normalized F/P weight ratio Group A (inappropriately light placenta with relatively heavy fetus) to normalized F/P weight ratio Group E (normal), there were significant differences in methylation in the following gene pathways: autism (adjusted p<0.0001), ADHD (adjusted p<0.001), epilepsy (adjusted p<0.01), and visuospatial dysfunction (adjusted p<0.05; Figure 3).

**Figure 3.**
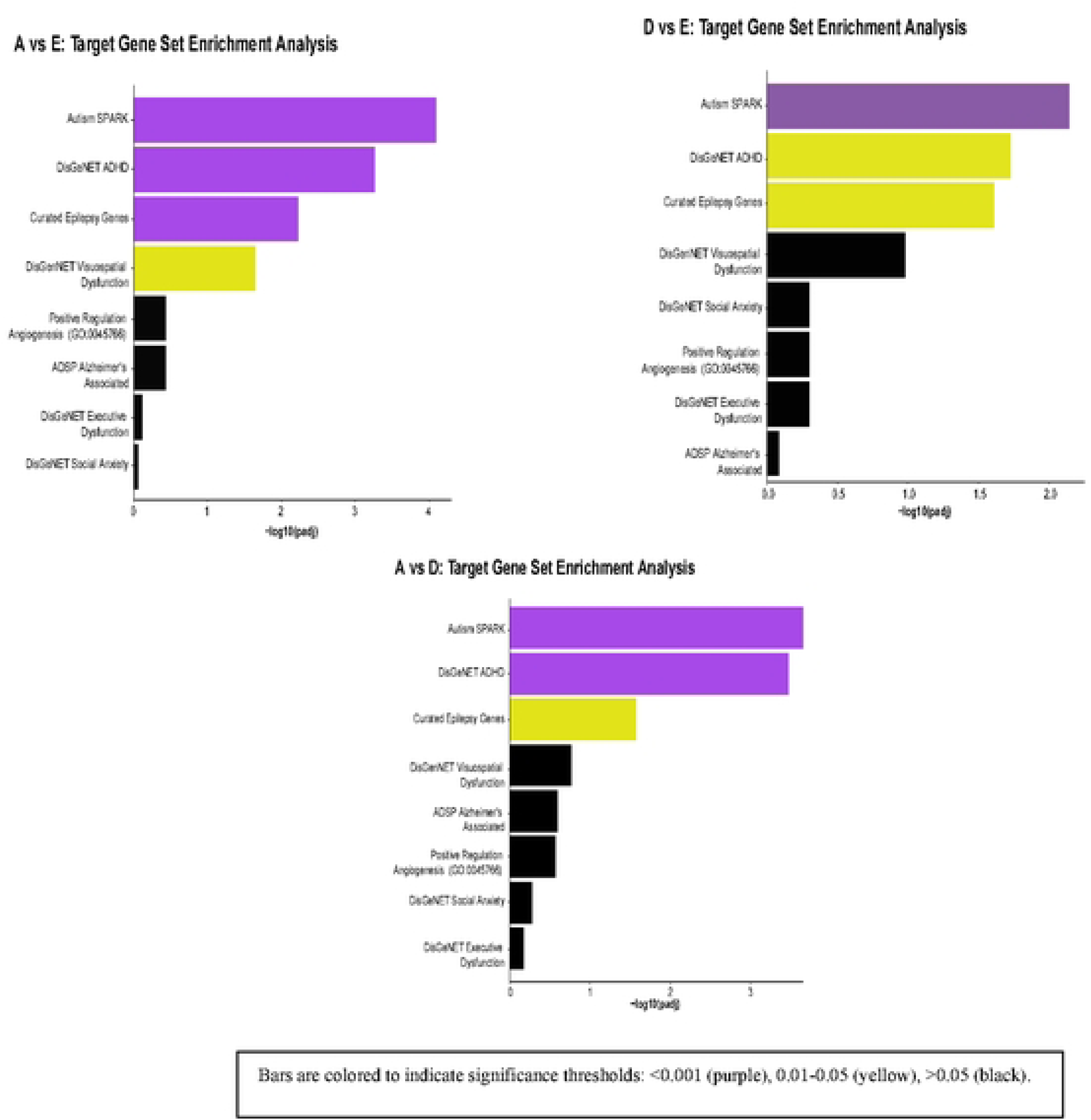
ND-associated Gene Pathways by Placental Ratio Groups, color-coded based on significance. We report p-value and Benjamini-Hochberg false discovery rate (FDR) adjusted p-values for each targeted gene set. Bars are colored to indicate significance thresholds: <0.001 (purple), 0.01-0.05 (yellow), >0.05 (black).

Comparison of normalized F/P weight ratio Group D (light placenta with relatively balanced growth of fetus) to normalized F/P weight ratio Group E (normal) similarly identified significant methylation differences in autism (adjusted p<0.01), ADHD (adjusted p<0.05), and epilepsy (adjusted p<0.05) gene pathway sets (Figure 3).

Significant methylation differences were also seen in autism (adjusted p<0.0001), ADHD (adjusted p<0.0001), and epilepsy (adjusted p<0.05) gene pathways when comparing normalized F/P weight ratio Group A (inappropriately light placenta with relatively heavy fetus) to normalized F/P weight ratio Group D (light placenta with relatively balanced growth of fetus; Figure 3).

### Placental Groups and BSID-III

Stratified by sex assigned at birth, for males, there were no significant differences between the normalized F/P weight ratio groups in the BSID-III scales of neurodevelopment for language (p = 0.549), motor (p=0.900), or cognition (p = 0.584; Figure 4a). For females, there were also no significant differences between the normalized F/P weight ratio groups in the BSID-III scales of neurodevelopment for language (p=0.543), motor (p=0.406), or cognition (p=0.719; Figure 4b).

**Figure 4.**
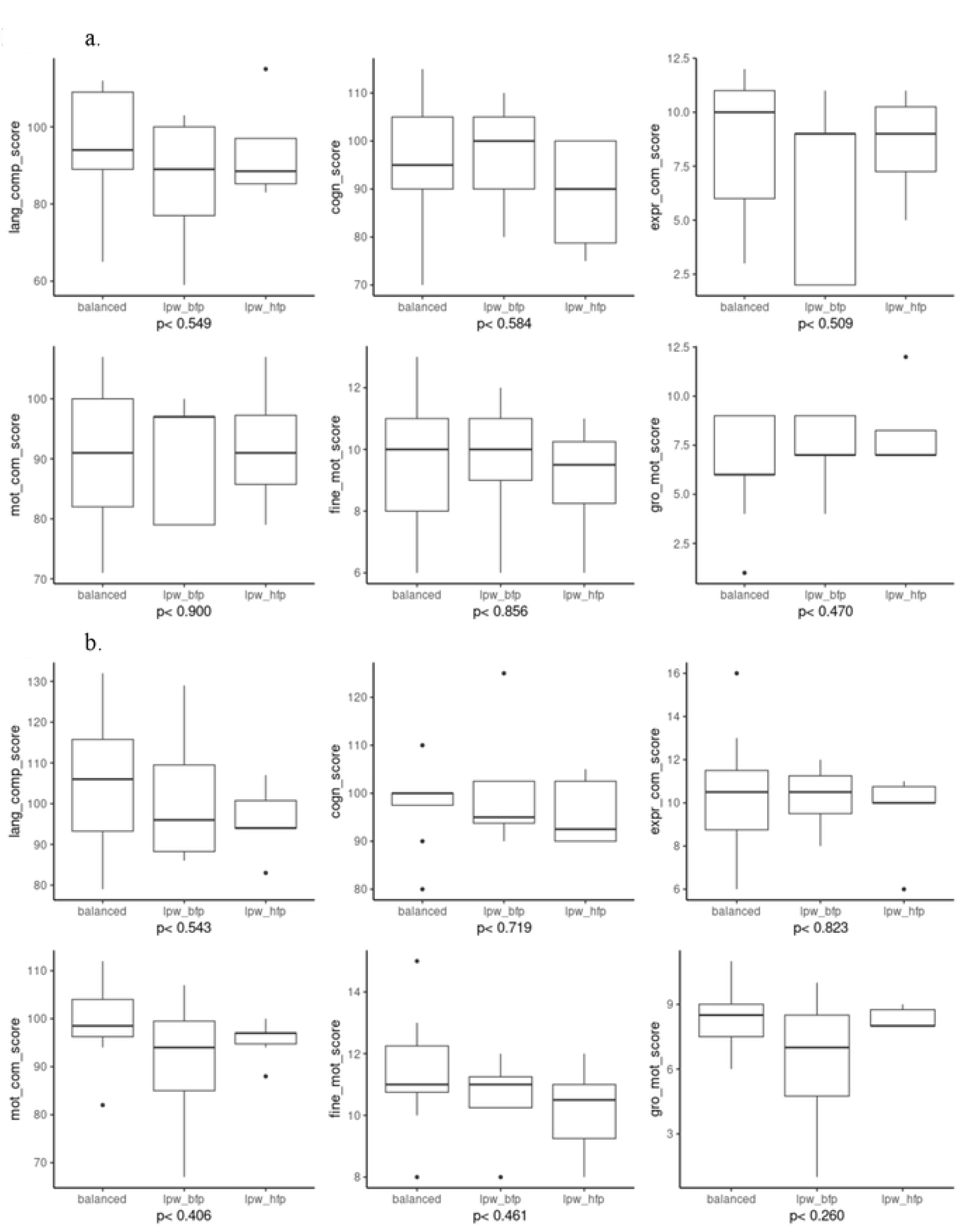
Box plot showing correlation of the Bayley Scales of Infant Developed (BSID-III) with F/P weight ratios categorized by sex (4a male, 4b female).

## Discussion

Infants with CHD demonstrated differentially methylated patterns of genes involved in nervous system development between normalized F/P weight ratio groups in support of F/P growth dynamics being relevant to neurodevelopmental outcomes in CHD.^16,55,56^ Furthermore, our data did not show significant methylation differences between normalized F/P weight ratios and BSID-III, but did identify differences in DNA methylation between normalized F/P weight ratio groups in gene pathways specific to common neurodevelopmental dysfunction in survivors of CHD. These results are in line with previous work demonstrating that abnormal placental pathology is related to perinatal, neonatal, and neurodevelopmental outcomes in both healthy pregnancies and pregnancies with CHD.^16,51,52^

Gene ontology enrichment analysis revealed a potential link between the placenta and neurodevelopment, with significant differential methylation identified between the normalized F/P weight ratio groups in multiple genes involved in various stages of neurogenesis. Notably, we uncovered this difference in peripheral methylation patterns by unbiased analysis. It is well known the importance of adequate nutrition and perfusion from the placenta to the growing fetus to ensure optimal neurodevelopment.^53^ Our data supports that abnormal placentation may influence pathways related to neurogenesis in the developing fetus. It is unclear how these peripheral changes in methylation correlate to potential disruption of the epigenome within the developing brain, though literature supports relevance between brain and peripheral signatures.^54,55^ Taken together, this data suggests that the peripheral epigenetic profile may be providing insight into pathways that contribute to altered brain development in CHD.

Many survivors of CHD have a spectrum of neurodevelopmental disabilities that become more apparent with age, including but not limited to executive dysfunction, visuospatial dysfunction, ADHD, autistic-like phenotypes, and social anxiety.^3,4^ As this population is now aging into adulthood, cognitive decline in early adulthood is being increasingly reported across all cardiac lesion phenotypes with features similar to Alzheimer’s disease.^24,56–59^ Early neurodevelopmental evaluation, both clinically and for research, is now most commonly with the BSID-III.^60,61^ Studies have suggested that the BSID-III may overestimate neurodevelopmental scores in survivors of CHD compared with prior versions of the neurodevelopmental scale.^60,61^ Categorized by sex, our data did not reveal significant methylation differences in BSID-III scores stratified by normalized F/P weight ratio groups. Given the above concerns about the BSID-III, the lack of differences in the BSID-III scores and the normalized F/P weight ratio groups in our cohort may be because this scale is insensitive for survivors of CHD, with poor correlation with long-term neurodevelopmental outcomes, as it is known that the majority of the CHD population has BSID scores that fall within 0.5-1 standard deviation of the normal population mean.^61^

In both human and animal models, there is increasing evidence that there are shared developmental pathways between the brain, heart, and placenta, including but not limited to angiogenic pathways, vascular endothelial growth factor (VEGF), and folic acid-associated pathways.^22^ However, there are still substantial gaps in understanding the relationship between the placenta, heart, and brain in CHD-affected pregnancies. Using previously reported gene set pathways for the above commonly reported neurologic dysfunction, significant differential methylation was identified, most notably, between the most unbalanced F/P weight ratio group and the balanced, normal F/P weight ratio group. This data suggests that there may be important epigenetic differences in the unbalanced fetal/placental interface when compared with balanced pregnancies in neonates with CHD.

This study is the first to report differences in DNA methylation using normalized F/P weight ratios in a cohort of neonates with CHD. Normalized F/P weight ratios have been used in other neonatal cohorts, but using these ratios to stratify groups for DNA methylation changes in CHD is novel. There are limitations that are important to acknowledge. First, recruitment for the parent study was not primarily for DNA methylation and required the exclusion of additional subjects to ensure appropriate methylation analyses. The small size of this cohort renders this study statistically underpowered. Our low statistical power and small sample size also required reduced stringency during multiple testing, increasing our risk of false positives. Second, this is a single-center study, and generalizability will be enhanced by data from other institutions or multi-center studies. Third, because of the small sample size of our cohort and the subsequent need to exclude subjects based on demographics, including race and ethnicity, we cannot generalize this to racially and ethnically diverse populations. Thus, this study provides intriguing avenues for longitudinal multi-institutional collaboration to understand if normalized F/P weight ratios and DNA methylation are biomarkers for neurodevelopmental disability in CHD.

## Conclusion

Gene ontology enrichment analysis revealed significant methylation differences in normalized F/P weight ratio groups, specifically in genes involved with neurogenesis. Furthermore, our data suggest that there may be notable DNA methylation differences between normalized F/P weight ratio groups in gene pathways implicated in neurodevelopmental disabilities that have been identified in aging survivors of CHD. Taken together, our data highlight the importance of further investigation into how DNA methylation changes in the placenta of fetuses with CHD impact postnatal neurodevelopmental outcomes, including if there is a direct epigenetic link between the placenta and neurogenesis in-utero.

## Data Availability

All relevant data are within the manuscript and supporting information files.

## Notes

### Competing Interest Statement

The authors have declared no competing interest.

### Author Declarations

The Children's Hospital of Philadelphia Institutional Review Board (IRB Approval Number: IRB 10-007823, IRB 16-013652)

